# Outcomes and Complications of Sickle Cell Disease among Hospitalized Paediatric Patients; A Retrospective Study in a Tertiary Hospital, Tanzania

**DOI:** 10.1101/2025.03.18.25324182

**Authors:** Adamu Kilungu, Ernest Winchislaus, Gaudence Alberto, Victor Kayombo, Abednego Wapalila, Tolbert Sonda

## Abstract

**Background:** Sickle cell disease (SCD) is a genetic blood disorder characterized by abnormal hemoglobin S, leading to various complications. This study aimed to assess the spectrum of SCD-related complications and outcomes among pediatric patients at Mbeya Zonal Referral Hospital in Tanzania.

**Methods:** A retrospective cross-sectional study was conducted, reviewing medical records of pediatric SCD patients admitted between June 2019 and June 2023.

**Results:** The study found an inpatient prevalence of 7.7% for SCD. Vaso-occlusive pain events (68%), infections (55.3%), and severe anaemia (27.7%) were the leading causes of admission. Low rates of hydroxyurea (11.4%) and penicillin V (28.3%) use was observed. The median haemoglobin level was 6.5 g/dL, indicating significant anaemia. Newly diagnosed patients (50%) had an average age of 5.12 years at diagnosis, suggesting delayed identification. The mortality rate was 3%.

**Conclusion:** These findings highlight the need of improved early diagnosis, management strategies, and access to essential medications for pediatric SCD patients in Tanzania. Implementation of newborn screening programs and increased awareness about SCD management could significantly improve patient outcomes.

## Introduction

Sickle cell disease (SCD) is a genetic blood disorder characterized by the production of abnormal haemoglobin, known as haemoglobin S (HbS). This abnormality causes red blood cells to become rigid and sickle-shaped under certain conditions, leading to impaired blood flow and various complications. The pathophysiology of SCD involves a point mutation in the HBB gene on chromosome 11, resulting in the substitution of glutamic acid with valine at the sixth position of the beta-globin chain. This single amino acid change promotes the polymerization of haemoglobin S under low oxygen conditions, causing red blood cells to sickle and obstruct blood vessels, leading to pain, organ damage, and increased risk of infections.

SCD is recognized as a significant global health concern, affecting approximately 5% of the world population. Annually, about 300,000 children are born with SCD, with 75% of these births occurring in Africa^1^. In Tanzania, around 14,000 babies are born with SCD each year, placing the country among those with the highest burden of the disease globally^2,3^. Without proper interventions, the prognosis for children with SCD in sub-Saharan Africa is dire, with high mortality rates before the age of five^4^.

Children with SCD are particularly vulnerable to severe complications such as acute chest syndrome, stroke, and infections^5–7^. Early childhood is a critical period for managing SCD to prevent these complications and improve long-term outcomes. However, many children in sub-Saharan Africa do not receive timely diagnosis or adequate care, exacerbating the disease’s impact^8^.

Comprehensive management of SCD is essential to improve patient outcomes and quality of life^9–12^. This includes early diagnosis, preferably through newborn screening programs, folic acid supplementation, penicillin prophylaxis, and the use of hydroxyurea. However, the implementation of these strategies in resource-limited settings like Tanzania remains challenging. Challenges in managing sickle cell disease in Tanzania include several critical issues. Firstly, the availability of hydroxyurea is limited, and even when it is available, the financial constraints of patients’ caretakers hinder access. Additionally, regulatory restrictions prevent general practitioners from prescribing hydroxyurea, limiting this authority to consultants only, as mandated by the major health insurance fund. Another significant barrier is the low awareness among clinicians regarding the benefits of essential medications like hydroxyurea and penicillin V prophylaxis in reducing complications associated with sickle cell disease^13,14^.

Furthermore, the absence of newborn screening programs leads to delayed diagnoses, contributing significantly to increased morbidity and mortality. Frequent hospitalizations, particularly in early childhood, are common due to vaso-occlusive pain events, infections, and severe anaemia, which are prevalent morbidities necessitating hospital admissions^3,14^.

This study aims to assess the spectrum of SCD-related complications and outcomes among pediatric patients at Mbeya Zonal Referral Hospital, providing valuable insights into the current state of SCD management in Tanzania.

## Methods

### Study Design and Setting

A retrospective cross-sectional study was conducted from January to March 2024 at Mbeya Zonal Referral Hospital (MZRH). MZRH is a tertiary healthcare facility located in Mbeya Urban, in the southern highlands of Tanzania. It serves as a referral centre for the surrounding regional referral hospitals in Mbeya, Njombe, Songwe, Ruvuma, Katavi, Rukwa and Iringa. The MZRH has a bed capacity of 826 and attends around 1200 outpatients and 120 inpatients. MZRH also functions as a teaching hospital, providing clinical training for medical students from the University of Dar es Salaam’s Mbeya College of Health and Allied Sciences.

### Data Collection

A comprehensive review of medical records of pediatric inpatients admitted between June 2019 and June 2023 was performed. Data pertaining to complications, administered treatments, prior utilization of hydroxyurea, folic acid, and penicillin V prophylaxis, demographic characteristics, and laboratory findings, including haematological profiles, were systematically recorded in case report form.

### Inclusion Criteria

The study included participants under the age of 18 who had been diagnosed with sickle cell disease and were admitted to the hospital between 2019 and 2023

## Data Analysis

Data was entered into Microsoft Excel and analyzed by using STATA version 16 (Stata Corp, College Station, Texas, USA). Descriptive statistics were summarized in frequencies and proportions (percent) for categorical variables, means with standard deviations or medians with interquartile ranges depending on the distribution of continuous variables were presented in tables and figures.

## Ethics

The study received ethical approval from the Mbeya Medical Research and Ethics Committee (Ref No: SZEC-2439/R.A/24/01) and permission from the Mbeya Zonal Referral Hospital administration. Researchers ensured confidentiality and privacy by excluding patient’s names and identifying information.

## RESULTS

### Prevalence and Demographics

The study identified 264 admissions (Fig. 1) with confirmed sickle cell disease (SCD) out of 3,424 total admissions, yielding a prevalence rate of 7.7%.

**Figure 1:**
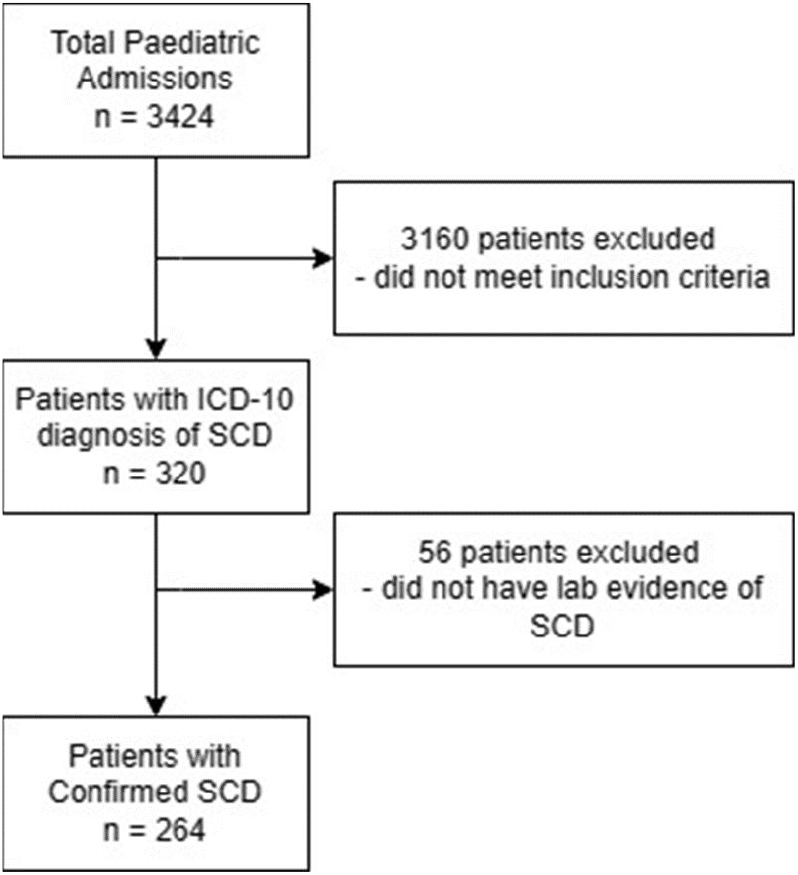
Flow diagram showing the selection of participants.

**Figure 2:**
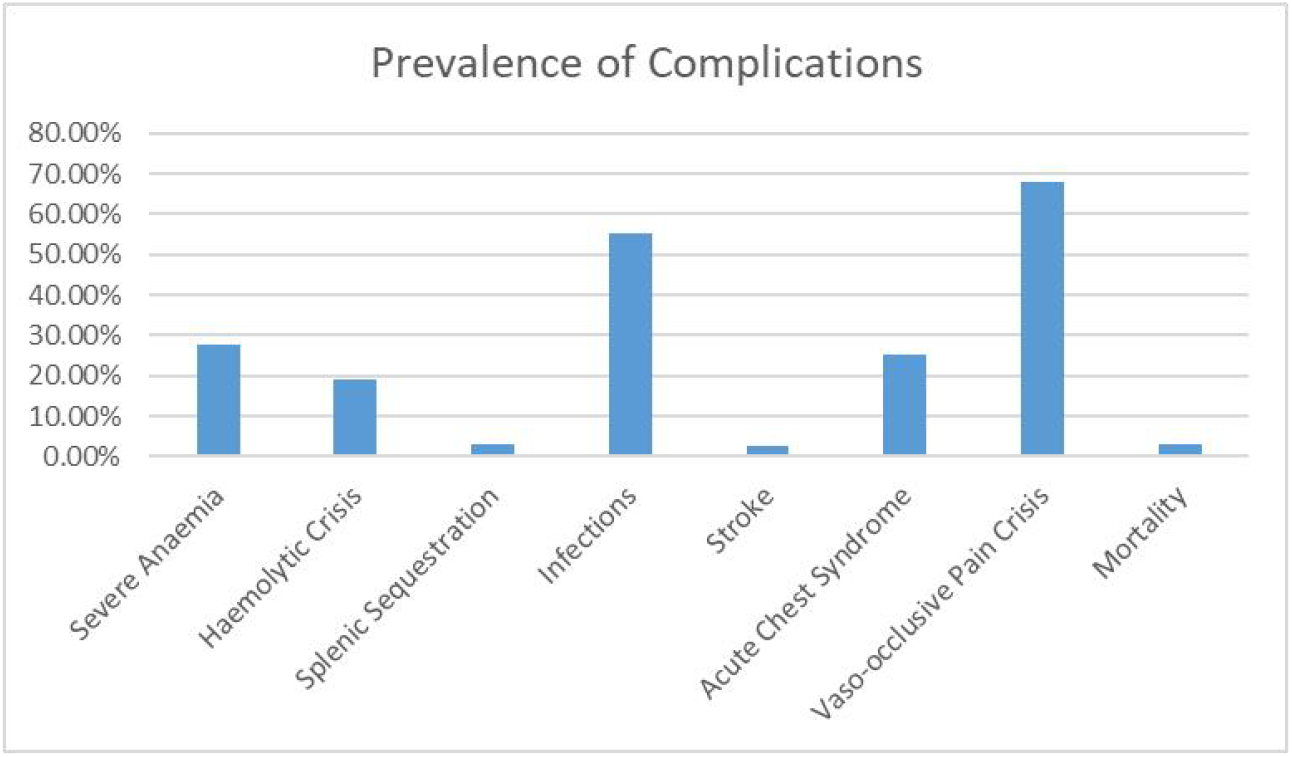
Prevalence of complications

The cohort exhibited a slight male predominance (53%, n=140). Age distribution analysis revealed that the majority of patients were young children, with 52.3% aged ≤5 years, 34.8% between 6-10 years, and 12.9% between 11-18 years.

Approximately, half of the participants (50.8%) had insurance coverage. Notably, 15% (n=40) of patients experienced multiple admissions (≥3), indicating a subset with recurrent hospitalizations. The average age at diagnosis for newly diagnosed SCD patients (n=132) was 5.12 years, suggesting delayed diagnosis in this population.

Regarding disease management prior to the index admission, 91% of participants reported the use of folic acid supplementation, while only 11.4% utilized hydroxyurea. Prophylactic penicillin V was used by 28.3% of patients. Additionally, chronic transfusion therapy, defined as receiving five or more blood transfusions, was administered to 11.4% (n=30) of patients (Table 1).

**Table 1:**
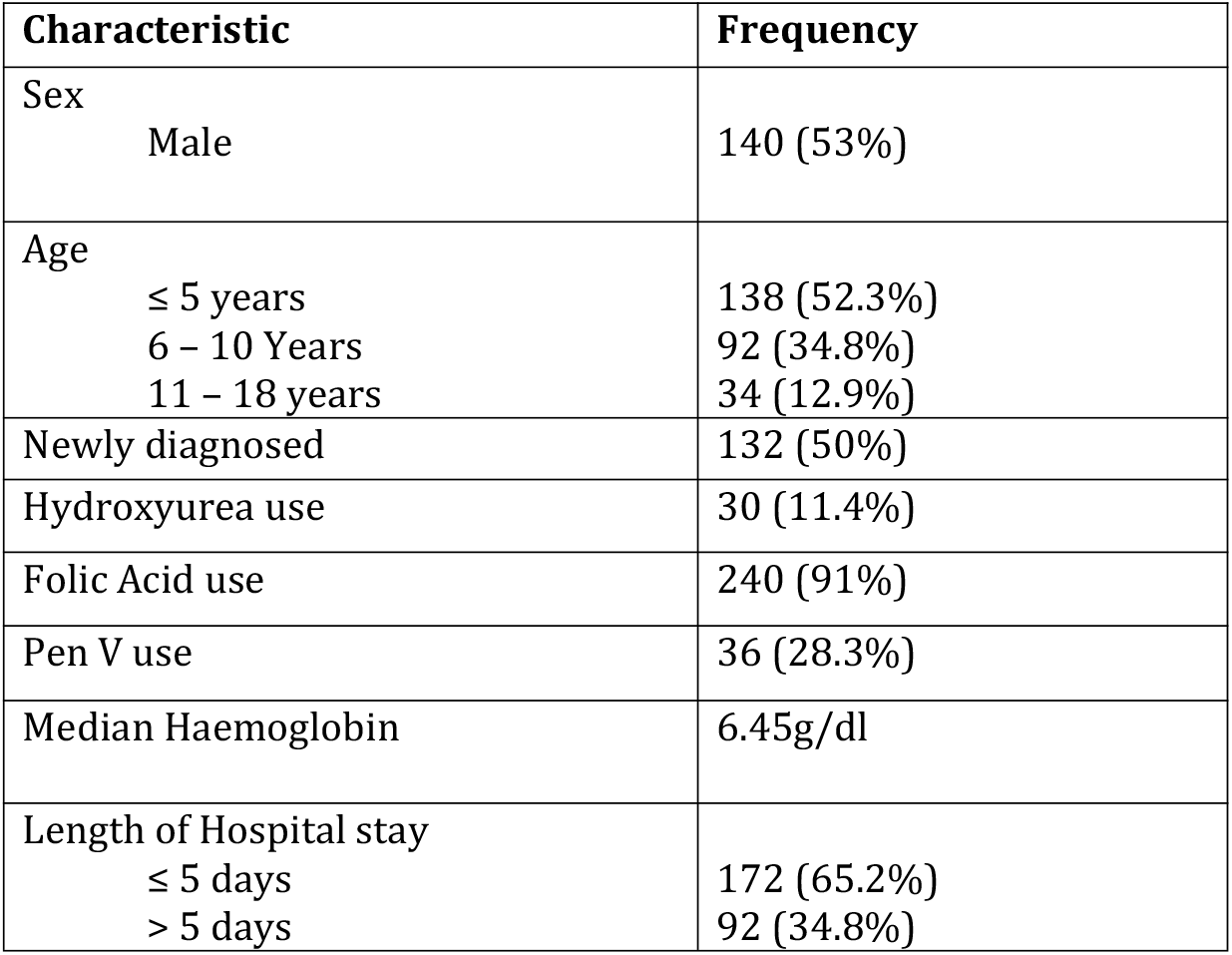
Patient historic characteristics.

**Table 2:** Haematological Profiles.

| Parameter (unit) | Median, IQR |
| --- | --- |
| Hb (g/dL) | 6.45 (4.75 – 7.75) |
| WBC ($\times 10^9/L$) | 18.9 (13.7 – 26.11) |
| Platelet count ($\times 10^9/L$) | 352 (224 – 469) |

**Table 3:** Information about blood transfusion.

| **NUMBER OF BLOOD**<br>**TRANSFUSION EPISODES** | **NUMBER OF**<br>**PATIENTS** | **PERCENTAGE**<br>**(%)** |
| --- | --- | --- |
| **0** | 157 | (59.5%) |
| **1** | 23 | (8.7%) |
| **2** | 23 | (8.7%) |
| **3** | 17 | (6.4%) |
| **4** | 13 | (4.9%) |
| **5** | 9 | (3.4%) |
| **>5** | 22 | (8.3%) |

### Haematological Profiles

The median haemoglobin concentration among the participants was 6.5 g/dL, indicating severe anaemia. Leukocytosis was observed in 79.9% of patients, with 83.8% exhibiting lymphocytosis and 62.9% showing neutrophilia. Platelet abnormalities were also noted, with thrombocytosis presented in 28.5% of patients and thrombocytopenia in 10%.

### Complications and outcomes

The study identified several complications and outcomes associated with SCD. Vaso-occlusive pain crisis was the most common complication, affecting 68% (n=179) of patients. Infections were reported in 55.3% (n=146) of cases, while severe anaemia was observed in 27.7% (n=73). Acute chest syndrome occurred in 25.4% (n=67) of patients, and hemolytic crisis was noted in 18.9% (n=50) of the patients. Other recorded complications include splenic sequestration in 3% (n=8) and stroke in 1.5% (n=4) of patients. The mortality rate among the study population was 3% (n=10). Regarding hospital stays, 65.2% of patients stayed five days or less, while 34.8% stayed more than five days.

## Discussion

The findings of this study, which was conducted at a tertiary facility in the southern highlands of Tanzania, reveal several critical aspects of sickle cell disease (SCD) management and its prevalence in pediatric patients. Key findings include a significant inpatient SCD prevalence, with vaso-occlusive pain events, infections, and severe anaemia identified as the top three causes of hospital admissions. The study also uncovered low utilization of essential SCD medications, including hydroxyurea and penicillin V prophylaxis. Additionally, the research highlighted the problem of delayed diagnosis, with many children being diagnosed at an age above five years.

The inpatient prevalence of SCD (7.7%) is notably higher than rates reported in similar studies from Malawi^6^ and Nigeria^15^. This discrepancy may be attributed to regional variations in SCD prevalence and differences in healthcare access and utilization patterns. The prevalence of complications, particularly vaso-occlusive pain events, infections, and severe anaemia, aligns with results from studies conducted in various regions^3,5,14,16–18^, underscoring the universality of these complications in pediatric SCD patients across different geographical and healthcare settings.

The study also found a median haemoglobin level of 6.5 g/dL, indicating significant anaemia in these patients. This mirrors findings from other research in Tanzania and across Africa^6,14,19,20^, where low haemoglobin values remain a common issue in SCD management. Additionally, the study showed that 40.5% of patients had a history of transfusions, raising concerns about potential complications like iron overload and alloimmunization, particularly in contexts with limited access to iron chelators and specialized blood products.

Another critical finding was the low utilization of essential SCD medications. Hydroxyurea was used by only 11.4% of patients, and 28.3% were on penicillin V prophylaxis. Despite some improvement compared to previous studies in Tanzania^14^, these rates remain suboptimal. Factors such as limited drug availability, financial constraints, misconceptions, and a lack of awareness contribute to this underutilization^13,19^. Addressing these issues through increased drug availability, financial support, and education for both healthcare providers and families is crucial for improving patient outcomes.

Lastly, the study highlighted the problem of delayed diagnosis and a relatively higher age at diagnosis among pediatric SCD patients. The findings showed that 52.3% of hospitalized children with SCD were below five years old, which is consistent with studies from other African countries^5,6,15^. Moreover, 50% of the children’s sickle cell status was unknown at the time of admission, with an average age at diagnosis of 5.12 years. This delay in diagnosis can be largely attributed to the lack of newborn screening programs in Tanzania, similar to many African countries^21^. Consequently, young children are more vulnerable to SCD complications, resulting in higher hospitalization rates. This consistency across the region emphasizes the urgent need for improved early diagnosis and management strategies for

## Conclusion

This study highlights the significant burden of sickle cell disease (SCD) among pediatric patients at a tertiary facility in Tanzania. Key findings include a high inpatient prevalence of SCD, with vaso-occlusive crises, infections, and severe anaemia being the predominant complications. Additionally, the study reveals concerning delays in SCD diagnosis and suboptimal utilization of essential medications like hydroxyurea and penicillin V.

## Strengths and Limitations

The study’s strengths include its large sample size, which provided a comprehensive view of SCD in a Tanzanian referral hospital setting. The inclusion of multiple aspects of SCD management and outcomes offered valuable insights into the current state of care for pediatric SCD patients in the region. However, the study had several limitations. Its retrospective design may have led to missing data or inaccuracies in medical records, potentially affecting the completeness and reliability of the findings. As a single-centre study, the results may not be fully generalizable to other regions or healthcare settings in Tanzania or sub-Saharan Africa.

## Data Availability

All data produced in the present study are available upon reasonable request to the authors

## Notes

### Competing Interest Statement

The authors have declared no competing interest.

### Funding Statement

This study was funded by Mbeya Zonal Referral Hospital

### Author Declarations

The study received ethical approval from the Mbeya Medical Research and Ethics Committee and permission

